# The motivational determinants of human action, their neural bases and functional impact in adolescents with OCD

**DOI:** 10.1101/2022.03.19.22272645

**Authors:** Iain E. Perkes, Richard W. Morris, Kristi R. Griffiths, Stephanie Quail, Felicity Waters, Margo O’Brien, Philip L. Hazell, Bernard W. Balleine

## Abstract

**Background:** Establishing the motivational influences on human action is essential for understanding choice and decision-making in health and disease. Here we used tests of value-based decision-making, manipulating both predicted and experienced reward values to assess the motivational control of goal-directed action in adolescents and the functional impact of OCD.

**Methods:** After instrumental training on a two action-two outcome probabilistic task, participants underwent Pavlovian conditioning using stimuli predicting either the instrumental outcomes, a third outcome or nothing. We then assessed fMRI during choice tests in which we varied predicted value, using specific and general Pavlovian-instrumental transfer (PIT), and experienced value, using outcome devaluation.

**Results:** Both predicted and experienced values influenced the performance of goal-directed actions in healthy adolescent participants, mediated by distinct orbitofrontal (OFC)-striatal circuits involving the lateral-OFC and medial-OFC respectively. To establish their functional significance, we tested a matched cohort of adolescents with obsessive-compulsive disorder (OCD). We found that choice between actions in OCD was insensitive to changes in both predicted and experienced values and that these impairments corresponded to hypoactivity activity in the lateral OFC and hyperactivity in medial OFC during specific PIT and hypoactivity in anterior prefrontal cortex, caudate nucleus and their connectivity in the devaluation test.

**Discussion:** We found, therefore, that predicted and experienced values exerted a potent influence on the performance of goal-directed actions in adolescents via distinct orbitofrontal- and prefrontal-striatal circuits. The influence of these motivational processes was severely blunted in OCD resulting in dysregulated action control associated with the intrusion of competing actions.

## Introduction

The capacity for goal-directed action allows us and other animals to control the environment in the service of our basic needs and desires(1,2). Such actions constitute, therefore, an adaptive and flexible form of behavioral control that depends on encoding the relationship between actions and their consequences, or outcome, during learning and on the value of the outcome for performance(3). Despite the functional significance of this capacity, the processes that generate the outcome values determining human action remain unclear(4,5). In contrast, considerable evidence from rodents suggests that value-based control involves two forms of incentive learning; one generated by stimuli that predict reward — called *predicted values(2,6,7)* — and a second induced by the direct experience of the emotional response evoked by contact with the specific goals or outcomes of goal-directed actions — called *experienced values*(6,8). These values influence instrumental performance in humans(9,10), and the broader circuitry that mediates this influence appears to be well conserved across species(11). Nevertheless, the way that these values are integrated with action-outcome retrieval during performance and the neural substrates that support that integrative process are underexplored in humans, as is their causal role in action control (12,13).

To examine these sources of motivational control, we developed behavioral tests that probe the influence of predicted and experienced values on human action(14–16). These forms of incentive learning have distinct psychological and behavioral determinants(17,18). However, recent evidence from rodents suggests that their influence on *performance* may involve the modulation of a final common pathway(19) involving orbitofrontal cortex (OFC)(20) and its projections to the striatum(21). In evaluating the neural bases of these incentive processes, therefore, we focused on this orbito-striatal circuit. Importantly, converging evidence from people with various psychiatric conditions suggests that aberrant orbitofrontal activity is associated with a range of symptoms, particularly those observed in obsessive-compulsive disorder (OCD)(16,22–24). To establish the functional significance of these incentive processes we compared their influence in healthy participants to a matched cohort diagnosed with OCD. Because longer duration of illness and its consequences often interferes with establishing the functional effects of psychiatric conditions (25), we focussed on a period early in the course of illness during adolescence using a sample of healthy adolescents as controls. If the OFC is critical for the motivational control of goal-directed action, then activity in the orbital-striatal circuit should relate to performance in healthy people whereas abnormal activity and connectivity should be predicted to attenuate value-based control of goal-directed action in people with OCD.

## Methods

**NOTE: The full unabridged methods – including participant instructions, data handling, data analysis and imaging procedures – is provided in the Supplementary Material**.

### Participants

21 healthy adolescents (control group) and 20 adolescents with a lifetime DSM-5 diagnosis of OCD (OCD group) were included in analysis. The sample size was based on power analysis drawn from a similar study(13) and stipulated a minimum sample size of n=16 to achieve 80% statistical power at an alpha of 0.05. There were no group demographic differences (**Table 1**). Consent or assent was provided by the participant, parent, or both. Inclusion and exclusions criteria, recruitment, diagnosis and pretesting are described in the **Supplementary methods – see also Table S1**. Participants with OCD were a representative sample; the mean Children’s Yale-Brown Obsessive Compulsive Scale score (CY-BOCS = 17; SD=9) was moderate, psychotropic medications were used by 70%, and 75% had a comorbid (lifetime) psychiatric diagnosis. People with OCD had greater symptoms of depression, anxiety and stress, however no individual scored higher than moderate. There were no significant group differences in age, gender, handedness, education, intelligence, hunger or food reward preference ratings (all *ts* < 1; see **Table 1**).

**Table 1.** Demographic and clinical characteristics

|  | *OCD*<br>(*n* = 20) | *Controls*<br>(*n* = 21) | *t*<br>(*df* = 38) | *p*-<br>*value* |
| --- | --- | --- | --- | --- |
| Age | 15<br>(2) | 15 (2) | 0.48 | 0.63 |
| Right handed (total) | 17 | 21 | 3.40 | 0.07 |
| Females (total) | 8 | 8 | 0.02 | 0.90 |
| English second language (total) | 2 | 5 | 1.73 | 0.19 |
| Years of education | 10<br>(2) | 11 (1) | 0.66 | 0.51 |
| Intelligence: WRAT | 48<br>(6) | 49 (5) | 0.56 | 0.58 |
| Symptom dimensions: DASS-21 |  |  |  |  |
| Depression | 7 (6) | 3 (4) | 2.53 | 0.02 |
| Anxiety | 7 (4) | 3 (3) | 2.98 | 0.01 |
| Stress | 12<br>(9) | 5 (4) | 2.92 | 0.01 |
| OCD diagnosis | 20 | 0 |  |  |
| OCD symptoms: CY-BOCS |  |  |  |  |
| Obsessions | 9 (5) | 0 |  |  |
| Compulsions | 8 (4) | 0 |  |  |
| Total | 17 (9) | 0 |  |  |
| Lifetime comorbid psychiatric diagnoses | 15 (75%) | 3 (14%) |  |  |
| anxiety disorder | 3 (15%) | 0 |  |  |
| depressive disorder | 9 (45%) | 0 |  |  |
| adjustment disorder | 2 (10%) | 2* |  |  |
| tic disorder | 4 (20%) | 0 |  |  |
| attention deficit hyperactivity disorder | 3 (15%) | 0 |  |  |
| substance use disorder | 2 (10%) | 0 |  |  |
| post-traumatic stress disorder | 1 (5%) | 0 |  |  |
| oppositional defiance disorder | 2 (10%) | 0 |  |  |
| eating disorder | 1 (5%) | 0 |  |  |
| elimination disorder | 1 (5%) | 1** |  |  |
| Current psychotropic medications | 14 (70%) | 0 |  |  |
| SSRI monotherapy | 5 (25%) | 0 |  |  |
| SSRI and anti-psychotic | 6 (30%) | 0 |  |  |
| other | 3 (15%) | 0 |  |  |

Notes: Means (SD or percentages) or totals of each demographic variable, along with symptom severity scores. Obsessive-compulsive disorder (OCD) Childrens Yale Brown Obsessive-Compulsive Scale (CYBOCS [score of 18 = moderate severity]), selective serotonin reuptake inhibitor (SSRI), Weschler Ranging Assessment Test (WRAT), Depression Anxiety Stres Scale 21-item version (DASS-21 [all means for cases in the moderate range; all means for controls in the normal range]), Children's Yale-Brown Obsessive Compulsive Scale (CY-BOCS). \*Adustment disorder with depressed mood in full sustained remission.
\*\*Encopresis, in remission. None of the control group had a current psychiatric disorder.

This study was approved by The University of Sydney Human Research Ethics Committee (2012/2284).

### Behavioral Methods

The battery of learning and performance tasks for human participants used here is based on tests developed and validated in rodents (**Figure 1**)(2,14). Participants abstained from eating for three hours prior to the experiment. Outcomes consisted of five different sweet or salty foods (see supplementary methods) and participants tasted and rated each food on a 7-point Likert scale (“Very Unpleasant” to “Very Pleasant”). Stimulus presentation and response recording was controlled by PsychoPy© software (v1.82.00) with responses recorded on a two-button response pad (Cedrus©, California).

**Figure 1.**
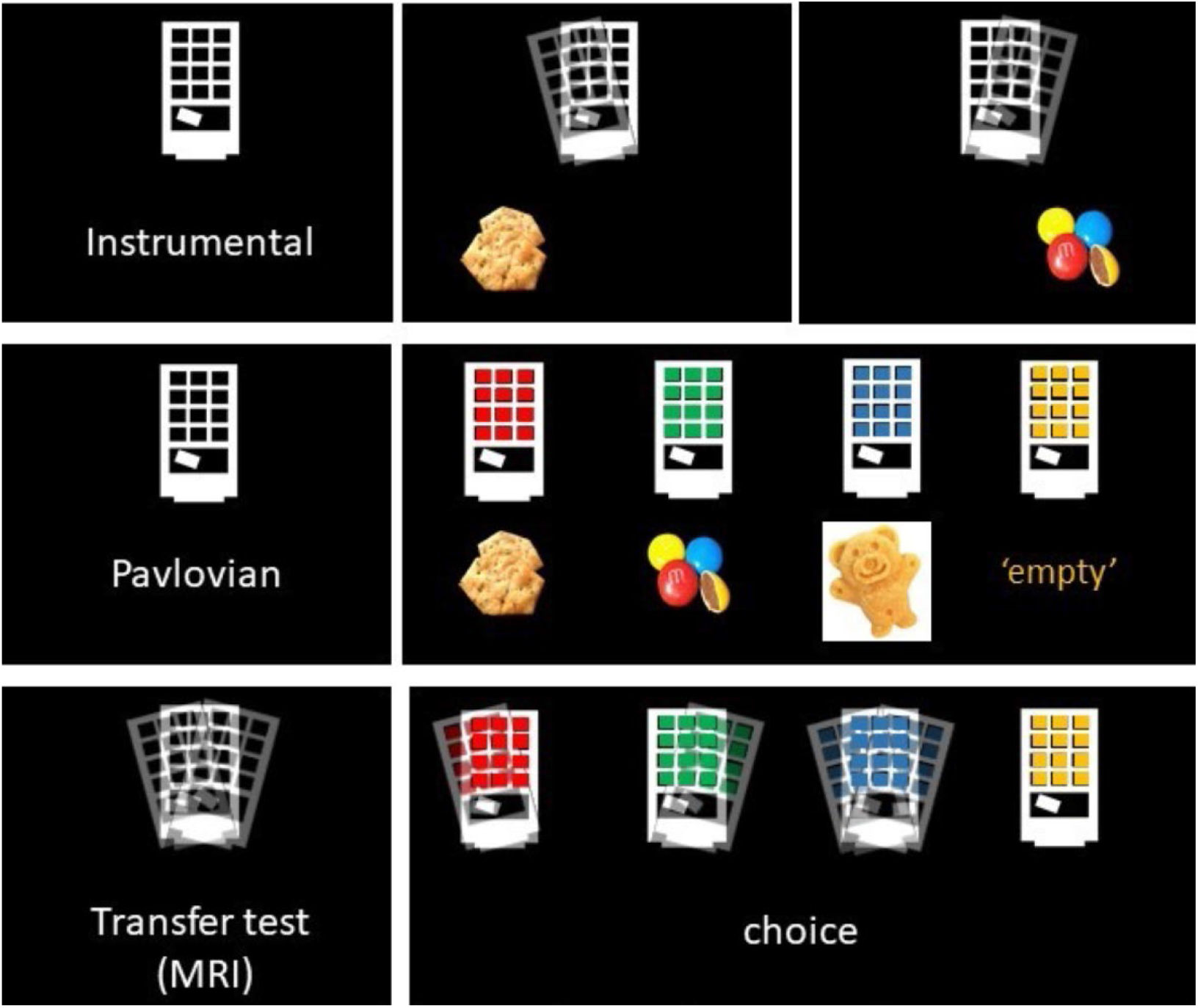
Experimental design for Pavlovian-instrumental transfer. **Top panels:** Instrumental conditioning was conducted by participants tilting a virtual vending machine to the left (A1) or right (A2) (actions) to earn food reward outcomes (O1, O2; shown on monitor for 1s) on a variable-ratio schedule (VR5). The task was continuous, not trial based, and participants could respond freely on each action until the end of each block – see Supplementary Methods for full details. **Middle panels:** Pavlovian conditioning involved four coloured lights (S1, S2, S3, S4) appearing on the machine for 6s in a random, non-replacing sequence within blocks which were followed by a multiple choice question. The stimuli predicted the appearance of an outcome (O1, O2, O3, O4(empty) counterbalanced) that occurred during the final 1-s of that stimulus. There then followed an intertrial interval followed by the next (randomly selected trial) – see Supplementary Methods for full details. **Lower panels:** During the Pavlovian-instrumental transfer test, the vending machine could again be freely tilted in either direct in a continuous manner both when it was unlit (i.e., during the 5-15s inter-trial intervals – it is - providing an active baseline measure), and when the coloured lights appeared on the machine. Four stimuli (S1, S2, S3, S4) appeared in a random, non-replacing sequence for 6s per stimulus every 18 seconds (0-4 second random jitter). Each stimulus was presented 12 times in random order. This transfer phase was conducted in extinction, i.e., no outcomes were delivered, so as to ensure that responding was not influenced outcome exposure on test – see Supplementary Methods for full details.

#### Instrumental conditioning

Left (A1) and right (A2) button presses were reinforced on a variable-ratio schedule (VR5) with a specific food, counterbalanced, dependent upon each participant’s three highest rated foods (**Figure 1; Supplementary Material**). The plate of snacks (O1) associated with A1 was placed on the desk on the left-hand side of the participant and the plate of snacks (O2) associated with A2 was placed on the right-hand side of the participant. As each outcome was earned, an image of that food appeared on the screen for 1-s and participants were invited to eat one piece of the relevant food. Participants were asked verbally which outcome was associated with which action. This phase ceased after a participant registered six consecutive correct answers.

#### Pavlovian conditioning

Prior to the start of conditioning the button box was removed. The three plates holding all three food rewards (O1, O2, O3) involved in Pavlovian conditioning were placed on the desk. Four stimuli (S1, S2, S3, S4) were paired with four outcomes (O1, O2, O3, Ø) (**Figure 1**). Two stimuli (S1, S2) were paired with two outcomes from instrumental conditioning, another stimulus (S3) was paired with an outcome (O3) not used in instrumental training and a fourth stimulus (S4) was paired with the word ‘EMPTY’ indicating no food was available. Stimuli were presented for 5 seconds, after which the image of the food outcome appeared beneath the stimuli for 1 second. The inter-trial-interval (ITI) was 10s (+/-5) during which neither stimuli nor outcomes were shown. After every block of four stimulus-outcome trials a multiple-choice question “Which snack will fall out?” appeared on the screen with a stimulus (coloured vending machine). If participants answered this question correctly they were invited to eat one piece of the relevant outcome. Pavlovian conditioning ceased after a participant registered six consecutive correct answers.

The next two phases were conducted in the MRI Scanner.

#### Pavlovian-instrumental transfer test (Figure 1)

For this test an MRI-safe button box was used. The four stimuli were presented individually for 6 seconds every 18 seconds (0-4 second random jitter). Each stimulus was presented 12 times in random order. Participants were able to tilt the vending machine during stimulus presentation and when the vending machine was unlit during the intertrial interval, providing an active baseline measure. This test was conducted in extinction, i.e., no outcomes were delivered, to ensure that responding was not influenced by the incidence of outcome delivery. PIT data for one participant from each group was lost due to error, leaving OCD *n*=19 and controls *n*=20.

#### Outcome devaluation procedure and test (**Figure 2**)

Participants were shown a 4-minute video of cockroaches crawling on one of the foods they had earned during instrumental conditioning (counterbalanced). The blank vending machine then appeared for 30 trials of 12 seconds each. Before each trial, a fixation cross was presented for 18 (±6) seconds. Participants could tilt the machine at any time. No outcomes were presented during the devaluation test. After the devaluation test, whilst still in the scanner, participants rated the desirability of O1 and O2 on a scale of Likert scale 1 to 7.

**Figure 2.**
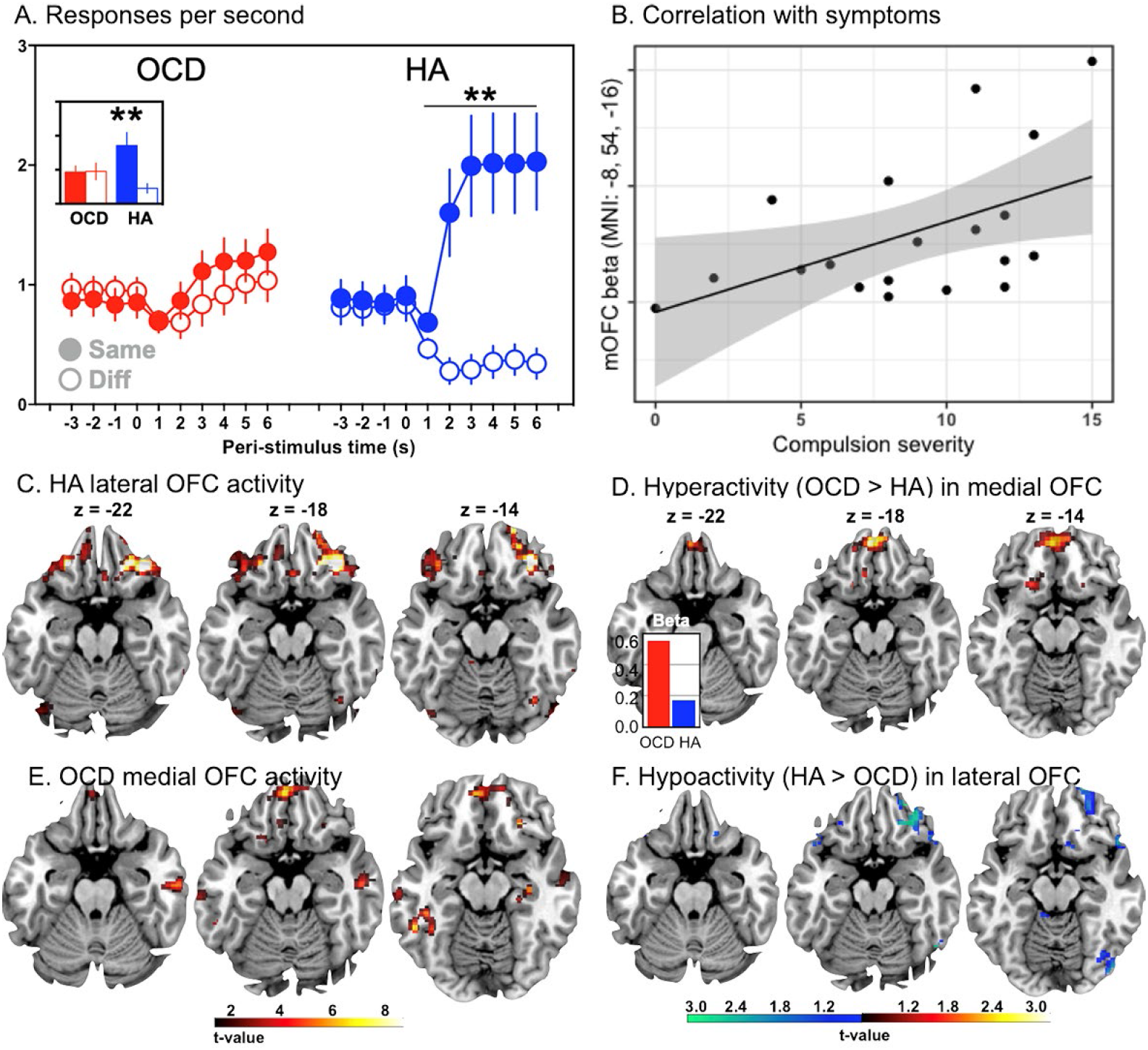
Predicted value — Specific Pavlovian-instrumental transfer test results. We tested whether participants could use outcome-specific predicted values, generated by stimuli S1 and S2, to direct choice toward actions associated with the ‘same’ outcomes as those predicted by the stimuli compared to actions associated with the outcome ‘different’ from that predicted (i.e., outcome-specific PIT). (**A**) Mean (±SEM) rates of button pressing per second before and during the specific stimuli when the action predicted the ‘same’ food reward as the stimulus, or a ‘different’ food reward from the stimulus. Post-hoc t-tests confirmed the action rate for the same food reward was significantly greater than the other action in healthy adolescents (HA) (*p* < .01). However, performance in the OCD group did not differ (p>0.05). The inset shows the mean performance across the test and illustrates the significant group x stimulus interaction (*p* =.017). (**B**) Adolescents with OCD showed hyperactivity in medial OFC relative to controls and the degree of hyperactivity correlated with compulsion severity. (**C**) Bilateral BOLD activity in the lateral OFC (right lateral OFC, BA47, MNI: 36,34,-18; left lateral OFC, BA47, MNI: -30,34,-20) tracked the effect of same vs. different stimulus on choice performance during the specific transfer test in healthy adolescents. (**D**) In contrast, relative to HA controls, OCD participants showed hyperactivity in medial OFC (BA11: MNI: -4, 50,-20) during the test, due to larger BOLD estimates in adolescents with OCD than negative BOLD estimates in those without (inset). (**E**) OCD participants also showed generally increased activity in medial OFC (BA11, MNI: -4,50,-20); however, (**F**) OCD participants showed significant hypoactivity in right lateral OFC relative to HA (BA47, MNI: 34, 34, -20).

After exiting the scanner participants re-completed the self-report hunger and food pleasantness scales presented at the start of behavioral training. They also completed a self-report six-item multiple-choice test of declarative recall of the instrumental (e.g., ‘What snack was associated with the LEFT key?’) and Pavlovian (e.g., ‘What snack was associated with the BLUE light?’) contingencies.

### Imaging methods

Scanning occurred in a 3T GE Discovery with a 32-channel head coil (GE Healthcare, UK). A T1-weighted high-resolution was acquired for each participant for registration and anatomical screening: 7200-msec repetition time; 2700-msec echo time; 176 slices in the sagittal plane; 1-mm slice thickness (no gap); 256-mm field of view; and 256 × 256 matrix. We acquired 300 T2*-weighted whole-brain echo planar images with a 2910-msec repetition time (TR); 20-msec echo time; 90-degree flip angle; 240-mm field of view; and 128 × 128 matrix with SENSE (Sensitivity Encoding). Each volume consisted of 52 axial slices (2-mm thick) with a 0.2-mm gap. Whole brain diffusion-weighted images were acquired using an echo planar imaging sequence with the following parameters: TR=8250ms; TE=85ms; number of slices=55 thickness=2mm-thick axial slices; matrix size, 128 × 128; in-plane resolution, 1.8 × 1.8mm^2^; 69 gradient directions. Eight images without gradient loading (B0 s.mm-2) were acquired prior to the acquisition of 69 images with uniform gradient loading (B0=1000s.mm-2).

### Data Analysis

The methods used to prepare apply statistical analyses to the behavioral, fMRI diffusion imaging and tractography data are fully detailed in the supplementary methods.

## Results

### Predicted values influence choice in healthy adolescents but not in those with OCD

There were no significant group differences in hunger rating (HA=5.9, OCD=6.2; t<1) or in the number of outcomes earned during instrumental conditioning (HA=18.5, OCD=18.7; t<1). Groups also did not have an action selection bias and showed a similar relationship between response rate and rating for preferred reward (Pearson r for cases and controls was 0.33 and 0.38, respectively). There were also no differences in Pavlovian conditioning. Healthy adolescents and those with OCD could recall these associations during training (HA=98%; OCD=91% t=1.39, p=0.1) after the choice tests and MRI scansMean group percentage correct was 98 and 93 percent, respectively (*p*=.40) (see Supplemental Materials for additional group comparisons).

In the specific PIT test (**Figure 1**)(7,26) the stimuli had different effects depending on the outcome they predicted (**Figure 2A**). Action rate was comparable between groups during the pre-stimulus (baseline) period (*t*_37_ < 1) – see the peri-stimulus periods prior to stimulus onset in **Figure 2A**. However, in HA, the specific stimuli (i.e., S1 and S2) elicited an immediate and potent elevation in the performance of the action that, during instrumental conditioning, resulted in the ‘same’ outcome as predicted by the stimulus, and a concomitant reduction in the performance of the action associated with the ‘different’ outcome. In contrast, the effect of predicted outcome values on choice was markedly impaired in adolescents with OCD who showed a mild but undifferentiated, that is general, increase above baseline for both the ‘same’ and ‘different’ actions during the specific stimuli (S1, S2). There was, therefore, clear evidence of significantly impaired specific transfer in OCD (planned group by action interaction *F*_1,37_=6.26, *p* =.017, 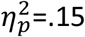).

We also tested the general arousing effect of predictive stimuli – i.e., General PIT(7) – by comparing responding during the generally rewarding (S3) and null (S4) predictors. Importantly, the general value prediction induced by S3 increased the performance of both actions (A1 and A2) relative to the null value prediction of S4 (**Figure 2B**) and, unlike the specific value predictions in the specific PIT test, this effect did not appear to differ between participant groups. The planned group-by-stimulus interaction conducted on action rates during the S3 and S4 stimuli was not significant (*p*=.47). There was, however, a significant main effect of stimulus (*F*_1,37_=7.22, *p* =.01, 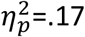).

### The influence of predicted values on choice is associated with orbitofrontal activation

BOLD activity was parametrically modulated by the rate of specific transfer during each trial of S1 and S2 (i.e., the rate on the ‘same’ minus ‘different’ action), revealed by the planned SPM t-test of healthy adolescents. The global peak voxels occurred in: (i) the bilateral OFC (right lateral OFC, BA47, MNI: 36,34,-18; t=7.35, k=122, pFDR=.025; left lateral OFC, BA47, MNI: -30,34,-20; t=6.05, k=14, pFDR=.026) **(Figure 2C)**; (ii) left dorsal caudate (Cd) (MNI: - 18,4,24; t=6.41, k=20, pFDR=.026); (iii) right putamen, fundus region (FPu) (MNI: 18,16,-2; t=6.14, k=20; pFDR=.026); (iv) the superior frontal gyrus (BA9, MNI: 8,56,34; *t*=5.84, k=12, pFDR=.032) and (v) the left supramarginal gyrus (SMG) in the inferior parietal cortex (BA40: MNI: -46,-30,36; t=7.26, k=138, pFDR=.025) **– refer Table 2 –** suggesting an orbito-striatal-parietal network actively modulated predicted value in healthy adolescents.

**Table 2.** Summary of the main findings

| Task | Group | MNI | Area | Descriptor | BOLD |
| --- | --- | --- | --- | --- | --- |
| Specific transfer | HA | 36,34,-18 | 47 | r. lateral OFC | ↑ |
| Specific transfer | HA | -22,34,-20 | 47 | l. lateral OFC | ↑ |
| Specific transfer | HA | -18,4,24 | – | l. lateral caudate nucleus (CdL) | ↑ |
| Specific transfer | HA | 18,16,-2 | – | r. Fundus of the putamen (FPu) | ↑ |
| Specific transfer | HA | 8,56,34 | 9 | r. dorsolateral prefrontal cortex (dlPFC) | ↑ |
| Specific transfer | HA | -46,-30,36 | 40 | l. inferior parietal lobe | ↑ |
| Specific transfer | OCD | -4,50,-20 | 11 | l. medial OFC | ↑ |
| Specific transfer | OCD | -58,-26,-18 | 21 | l. middle temporal gyrus (MTG) | ↑ |
| Specific transfer | OCD | 58,-18,-24 | 21 | r. middle temporal gyrus (MTG) | ↑ |
| Specific transfer | OCD | 44, 32, -8 | 47 | r. lateral OFC | ↓ |
| Specific transfer | OCD | -18,2,26 | – | l. dorsal caudate | ↓ |
| Specific transfer | OCD | 34, 34, -20 | 47 | r. lateral OFC | ↓ |
| Specific transfer | OCD | 20,18,0 | – | r. putamen | ↓ |
| Specific transfer | OCD | 6,58,32 | 9 | r. dorsolateral prefrontal cortex (dlPFC) | ↓ |
| Specific transfer | OCD | -8,54,-16 | 10 | l. medial OFC | ↑ |
| General transfer | BOTH | 12,38,-6 | 11 | r. medial OFC, posterior | ↑ |
| Outcome deval | HA | -14,6,18 | – | l. medial caudate nucleus (CdM) | ↑ |
| Outcome deval | HA | -22,60,2 | 10 | l. anterior PFC | ↑ |
| Outcome deval | HA | -18,30,-2 | 13 | l. centrolateral OFC | ↑ |
| Outcome deval | HA | 14,54,-2 | 10 | r. anterior PFC | ↑ |
| Outcome deval | OCD | 2,50,10 | 10/32 | r. anterior PFC/dorsal ACC | ↓ |
| Outcome deval | OCD | 6,18,-6 | 14 | r. anterior PFC–caudate tract | ↓ |

In contrast, adolescents with OCD showed localised hyperactive neural responses associated with outcome-specific predictions compared with healthy adolescents in the planned whole-brain SPM t-test. The greatest hyperactivity occurred in the left medial orbital gyrus (MOrG)/medial OFC (BA11: MNI: -4, 50,-20; *F*_2,37_=31.33, k=582, pFDR < .001) (**Figure 2D)**. Examination of the BOLD parameter estimates for each group confirmed that this difference was due to larger BOLD estimates in adolescents with OCD rather than negative BOLD estimates in those without (**Figure 2D inset**). No between-group differences in activity in the caudate or cingulate cortex were found; however there were significant differences in the right lateral OFC (BA47, MNI: 44, 32, -8, F = 29.69, k = 516, pFDR < .001), left and right middle temporal gyri (e.g., BA21, MNI: -58,-26,-18; *F*=22.95, k=693, pFDR<.001; BA21, MNI: 58, -18, -24, F = 22.91, k = 259, pFDR < .001) – **Table 2**. To explore the potential source of the BOLD hyperactivity in OCD, we conducted follow-up ROI analyses centred on the global peak voxels from the specific transfer test of the healthy adults using a 4 × 4 × 4 mm search space in each case. Hypoactivity (relative to HA) was found in the dorsal caudate (MNI: -18,2,26, t=4.44, pFWE < .001), right lateral OFC (BA47, MNI: 34, 34, -20; t=2.18, pFWE = .048), but not the left lateral OFC (pFWE = .35) – refer **Figure 2F**. Posthoc ROI tests also revealed hypoactivity in the right putamen (MNI: 20,18,0; t=2.45, pFWE=.028), and superior frontal gyrus (BA9: MNI: 6,58,32; t=2.8, pFWE=.01) (See Supplementary Results for covariate analyses for age, WRAT or handedness and Supplementary Results and **Figures S1A, S1B & S1C** for tractography results).

Finally, we assessed the relationship between compulsion symptoms and the largest hyperactive BOLD response in the medial OFC in the OCD adolescents in the follow-up ROI correlation analysis (**Figure 2B**). This analysis included obsession and compulsion severity as SPM covariates-of-interest and revealed a significant positive correlation between compulsion severity and hyperactive BOLD responses in the left medial orbital gyrus (MOrG)/medial OFC (BA11: MNI: -8,54,-16; t=2.79, pFWE=.047, svc) suggesting medial OFC hyperactivity may be related to the hypoactivity observed in lateral OFC.

### General PIT produced activity in a posterior region of medial OFC

In General PIT no significant differences between groups in action rate during the general reward stimulus (S3) and the null reward stimulus (S4) were found (**Figure 3A**) and so we collapsed this measure across groups and examined the effect of incentive motivation on neural activity in a whole-brain analysis. A posterior region of medial OFC activity tracked changes in the predicted values across presentations of the general predictive stimuli (S3 and S4) in both groups (**Figure 3B**: BA11: MNI: 12,38,-6; *t*_*38*_=4.71, k=67, pFDR=.044), indicating that the influence of general reward arousal on brain activity was generally intact in healthy adolescents and those with OCD. There were no significant group differences in the follow-up ROI comparison (pFWE = .19), indicating that, on average, the influence of general reward predictions on this region of medial OFC was similar in the two groups.

**Figure 3.**
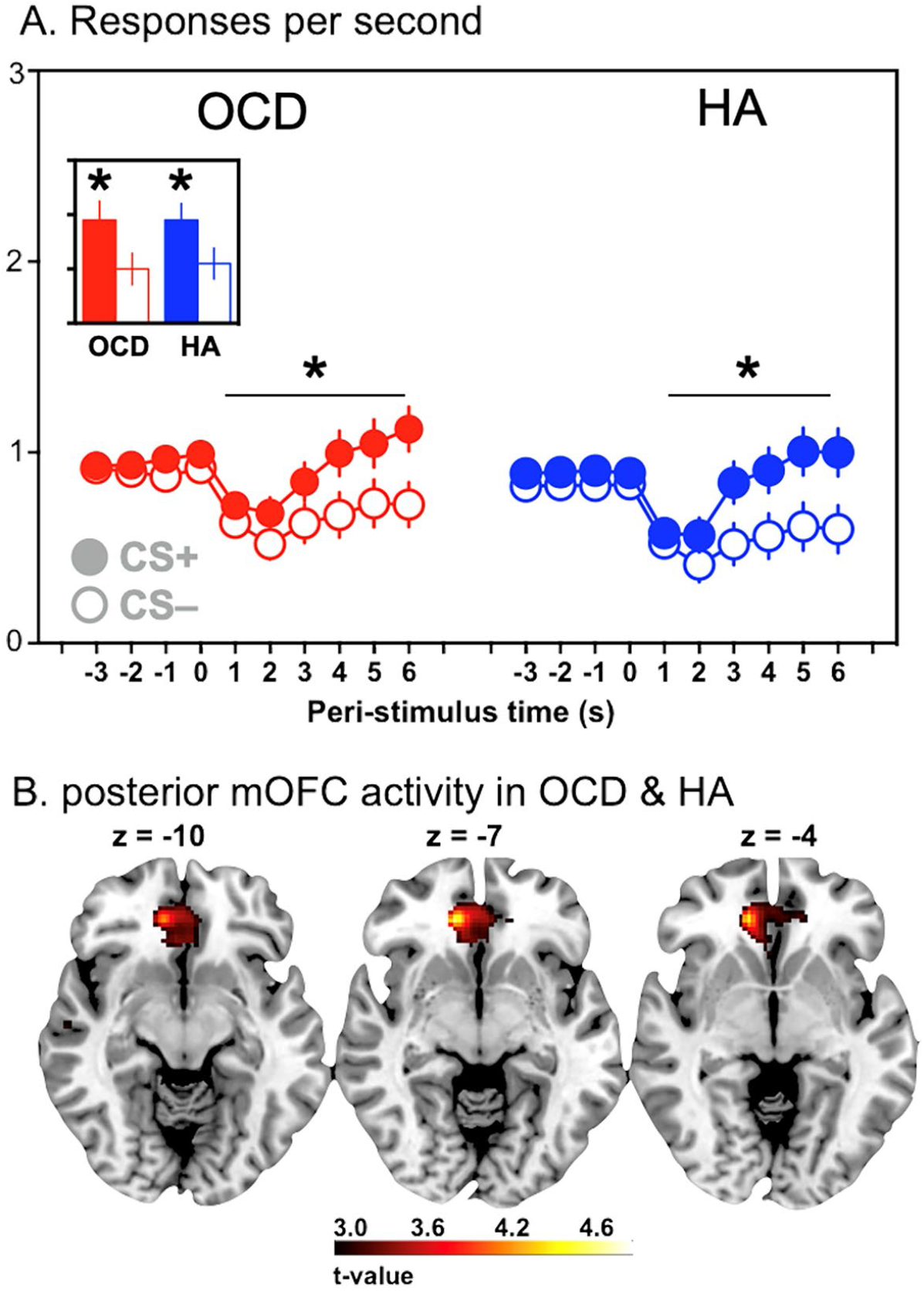
Predicted value — General Pavlovian-instrumental transfer test results. (**A**) Mean (±SEM) response rates (per second) before and during the general excitatory stimulus, S3 associated with the reward that was not presented during instrumental training, and the neutral S4 stimulus associated with the ‘empty’ vending machine. Post hoc t-tests confirmed the response rate during S3 was significantly greater than S4 in both HA and OCD groups, *ps* < .05. (**B**) Activity in a posterior region in the medial OFC (BA11: MNI: 12,38,-6) tracked changes in the predicted values across presentations of the general predictive stimulus (S3) in both groups.

### The effect of experienced value on choice

Next, we manipulated experienced value to test its influence on choice in an outcome devaluation test. To induce devaluation, participants watched a short video showing cockroach infestation of one of the two outcomes (O1 or O2) used in instrumental conditioning. They were then given a choice test on the vending machine in nominal extinction, i.e., no coloured lights were shown and no snacks seen or consumed – **Figure 4**.

**Figure 4.**
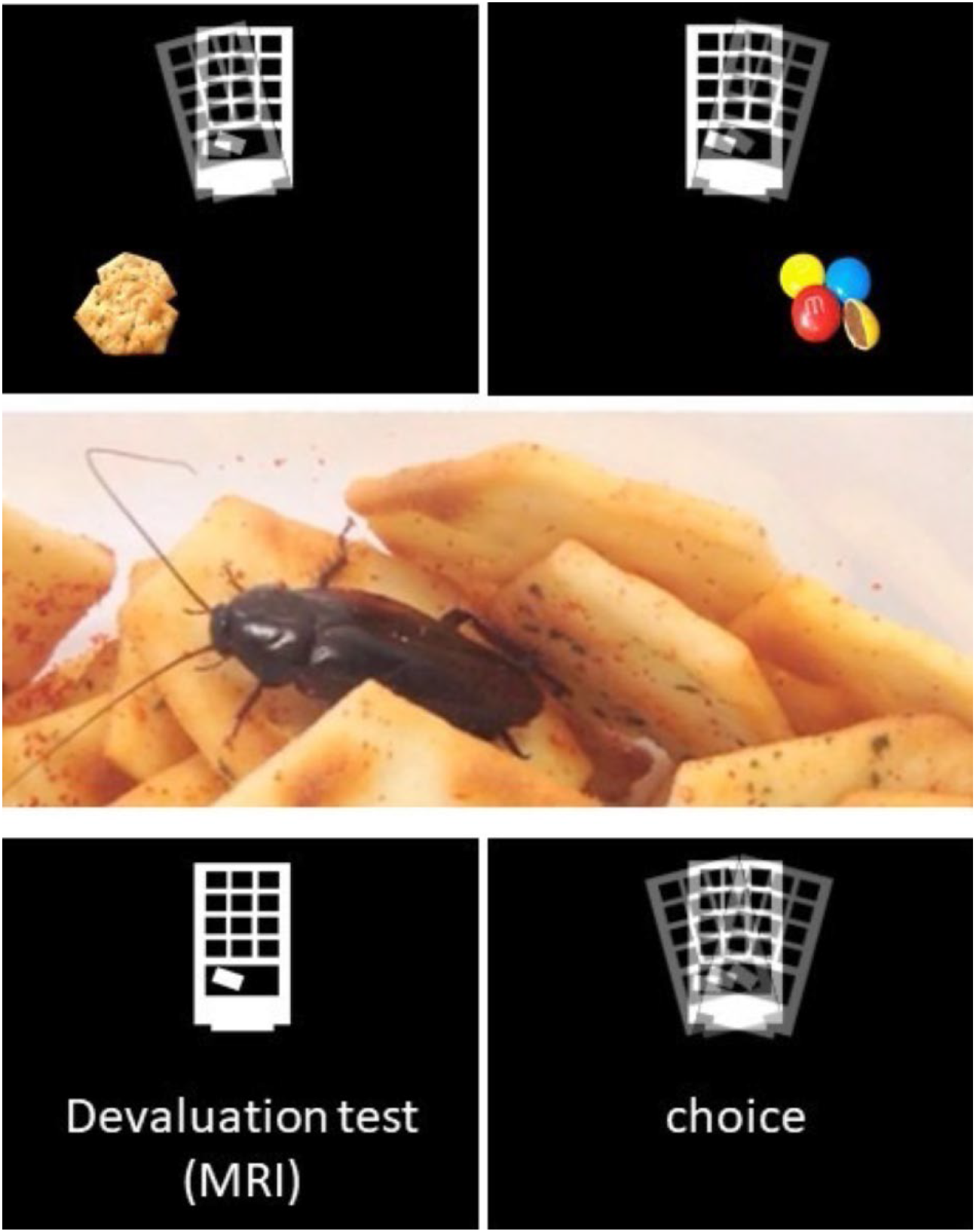
Outcome devaluation. After instrumental training (e.g., as in top panels) participants watched a video showing one of the outcomes infested with cockroaches as a devaluation treatment (middle panel). This treatment was effective (see Figure 5B). After the video we conducted a test in which all participants could tilt the vending machine to the left or right allowing us to assess their choice performance. Importantly, the devaluation test was conducted in extinction; i.e., no outcomes were delivered during the test.

After viewing the video, healthy adolescents preferred the action previously associated with the ‘valued’ outcome, i.e., the outcome that had not been ‘devalued’. In contrast, people with OCD showed no behavioral preference between A1 and A2 responding similarly on the valued and devalued actions (**Figure 5A**). 3-way ANOVA (group x devaluation x time) confirmed the interaction between group and devaluation was significant (*F*_1,185_=8.028, *p*=.005, 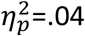): healthy adolescents responded significantly more on valued than the devalued action (t = 2.84; df = 37, p = .007) whereas OCD participants did not (t = 1.14; df = 35, p = .26) (**Figure 5A inset**). To satisfy fMRI protocols the outcome devaluation test was 15 min duration and there was a main effect of time as responding decreased across the test (*F*_4,148_ = 3.45, *p*<.001, 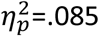) but no group x action x time interaction (*F*_4,185_ = 0.11, *p*=.979, 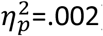). Simple effects analysis of the significant two-way interaction revealed a difference in performance between groups on the valued action (t = 1.41, df = 36, p = .005) but not devalued action (t = 0.68, df = 34, p = 0.49). As participants were instructed to use one finger during the training and tests, this reduced performance on the valued action likely reflects response competition.

**Figure 5.**
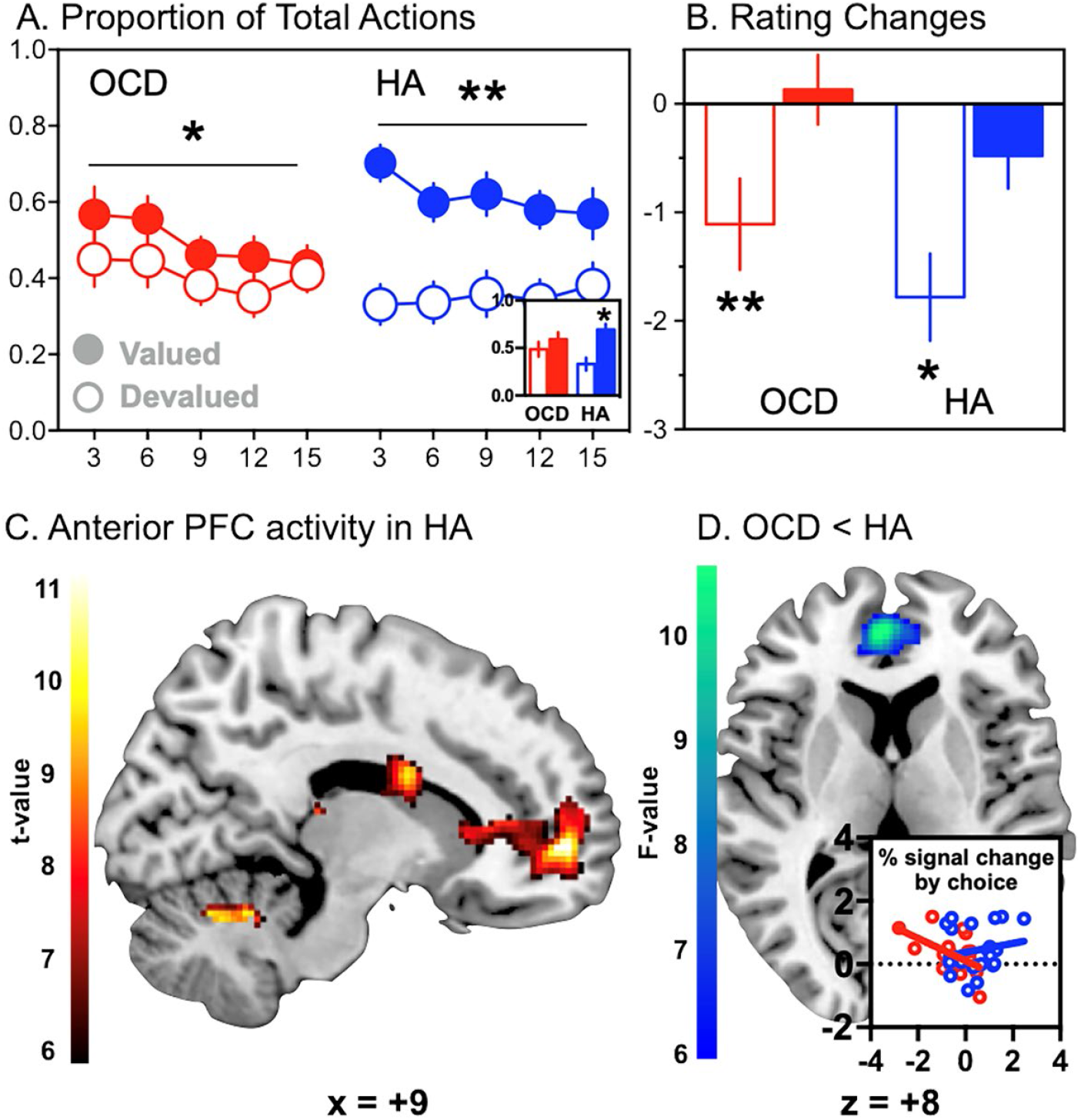
Experienced value — Devaluation test results. (**A**) Proportion of total button presses on the actions that previously delivered the ‘valued’ and the ‘devalued’ food rewards during each minute of the devaluation test in 3 min blocks. After viewing the outcome devaluation video, healthy adolescents preferred the action previously associated with the still ‘valued’ (non-contaminated) outcome vs. the now ‘devalued’ (contaminated) outcome. In contrast, people with OCD showed no behavioral preference responding similarly on the valued and devalued actions. Inset illustrates the significant interaction in the average performance across the session (*p*=0.005) (B) Mean (±SEM) change in the food ratings of the devalued (open bar) and non-devalued food (filled bar) after devaluation (pre – post). (**C**) BOLD activity tracked valued actions in the left dorsal caudate (MNI: -14,6,18), an anterior region of anterior prefrontal cortex (BA10, MNI: -22,60,2), medial orbital gyrus (MOrG)/centrolateral OFC (BA13, MNI: -18,30) and an anterior rion of the medial OFC (BA10: MNI: 14,54,-2). (**D**) Group differences were detected in the anterior PFC revealing reduced activity in the right anterior prefrontal cortex extending into the right dorsal anterior cingulate (dACC; BA32) of the adolescents with OCD (BA32: MNI: 2,50,10). (**D - Inset**) The inset shows the correlation between activity at the peak voxel per participant correlated with a devaluation score calculated as the average response rate on the valued action minus the average rate on the devalued action. OCD group in red (*r*=–.51), HA group in blue (r=+.17).

Interestingly, food desirability ratings showed no inter-group differences in the change in outcome desirabilty ratings (post–pre) for the food outcomes – **Figure 5B**. Two-way ANOVA revealed a significant main effect of devaluation on food ratings (*F*_1,39_ =24.38, *p* < .001, 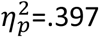) but neither a main effect of group nor a significant interaction (interaction *F*_1,39_=0.015, *p*=.903, 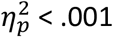). Despite the difference in choice performance, the correlation between outcome rating and action rate for the valued vs. devalued outcome was significant in both groups (*r*=.604 and .628, *p*s < .05; OCD and HA, respectively). Linear regression confirmed there was no significant difference in slope between groups (*p*=.96). Therefore, although choice performance after outcome devaluation clearly differed between groups, there was no difference in sensitivity to the devaluation treatment *per se*.

### Anterior PFC and dorsal caudate activity tracked the effect of experienced value on choice

BOLD activity that tracked choice driven by experienced value was determined by the planned SPM t-test in healthy adolescents: significant effects occurred in the left dorsal caudate (MNI: -14,6,18; *t*_19_ = 17.75, pFDR < .001), an anterior region of left middle frontopolar gyrus (MFPG)/ anterior prefrontal cortex (BA10, MNI: -22,60,2; *t*_19_ = 16.44, pFDR < .001), medial orbital gyrus (MOrG)/centrolateral OFC (BA13, MNI: -18,30,-2; *t*_19_ = 14.76, pFDR < .001) and an anterior region in the right middle frontopolar gyrus (MFPG)/medial OFC (BA10: MNI: 14,54,-2; *t*_19_ = 13.19, pFDR < .001) shown in **Figure 5C – refer Table 2**. Group differences were detected within the follow-up ROI analysis of this cluster within the anterior PFC region (OMPFC)(27) and revealed a significant deficit in activity in the right anterior prefrontal cortex (AntPFC; BA10) extending into right dorsal anterior cingulate (dACC; BA32) of the adolescents with OCD (**Figure 5D**, BA32: MNI: 2,50,10; *F*_2,39_ = 10.64, k = 29, pFWE = .028, svc).

Parameter estimates from the peak voxel in this ROI were extracted per participant and correlated with a devaluation score calculated for each participant, i.e., the average response rate on the valued action minus the average rate on the devalued action. The correlation between BOLD activity and this devaluation difference score was *r*=–.51 and +.17 in people with and without OCD, respectively, and these relationships differed between groups, p=.046 (**Figure 5D inset**), suggesting that, in adolescents with OCD, hypoactivity in this anterior PFC/dorsal ACC region resulted in a deficit in goal-directed choice after a change in experienced value.

### Anterior PFC-caudate tract strength is weaker in adolescents with OCD

Using the anterior PFC/dorsal ACC region identified above as a seed region, tractography analysis compared afferent and efferent tracts between groups to investigate neural disconnection as a contributing factor to task performance. This analysis revealed a lower tract strength in the projection to the caudate nucleus (peak MNI coordinates: 6,18,-6; cluster size: 56 voxels) in adolescents with OCD relative to healthy controls (pFWE=.008; **Supplementary Figure S2A**). **Supplementary Figure S2B** shows the average connectivity, across all participants, of the anterior PFC/dorsal ACC seed mask (top 0.02% of tracts sent from this seed mask).

## Discussion

We assessed the role of the primary incentive processes controlling goal-directed action in adolescents: reward values derived from Pavlovian predictions, or *predicted values*, and those derived from direct, largely consummatory, contact with the goal of goal-directed actions, or *experienced values*(2,6,17). We found that: (i) in specific PIT, stimuli generating specific reward predictions biased choice towards actions earning the outcome predicted by the stimulus, (ii) in general PIT, general reward predictions elevated the performance of actions regardless of outcome and (iii) in outcome devaluation, changes in experienced value biased choice of action away from the devalued action and towards that associated with the still-valued outcome.

### The neural bases of motivational control

Using fMRI, we found considerable commonality in the neural determinants of these incentive processes with those documented in rodents but with some potentially important differences. For example, the specific reward predictions in the Specific-PIT test induced increased activity bilaterally in the lateral OFC, right dorsolateral PFC, right putamen, left medial OFC, left lateral caudate, and left inferior parietal lobe. In contrast, although earlier work implicated lateral OFC and dorsal striatum(29,32,33), it also implicated circuits involving ventral striatum, including the accumbens shell and ventral pallidum in rats(30,31), ventrolateral putamen in humans(32), and basolateral amygdala in both species(33,34). As such, the orbito-striatal-parietal circuit observed here may suggest an important target for future work investigating how predicted values affect performance. Similarly, we found that the influence of general reward predictions in the General-PIT test increased activity in a posterior region of the right medial OFC, as we have found previously(14), whereas prior work focused on a circuit involving central amygdala and accumbens core in both rodents and humans(34–37). This work suggests medial OFC might be added to this circuit and, indeed, it maintains a strong relationship with ventral striatum in a number of potentially related functions(38). Finally, the influence of expected reward values on choice following outcome devaluation engaged a circuit linking an anterior part of both the left and right prefrontal cortex with the medial caudate, heavily implicated in previous anatomical and functional work(19-23). In addition, as we found previously in assessing the calculation of action values(39,40), we also found evidence of dorsolateral PFC involvement.

### The functional significance of this circuitry as revealed by OCD

In contrast to healthy adolescents, we found that, behaviorally, the influence of both forms of incentive process on choice performance was blunted or abolished in OCD. In specific-PIT OCD participants showed a mild general elevation in the performance of both actions irrespective of the outcome predicted by the stimuli and, in the outcome devaluation test, OCD participants responded similarly on the valued and devalued actions. Nevertheless, on other measures, OCD participants did not differ from controls. For example, our assessment of their Pavlovian and instrumental conditioning suggested they learned the specific stimulus-outcome and action-outcome associations similarly to controls and showed similar changes in outcome desirability ratings after the devaluation treatment. Of course, it is possible the measures of conditioning and of changes in value that we used were less sensitive than the specific transfer and devaluation tests in detecting group differences. It is also possible that the verbal assessment we used to check our participants’ ability to retrieve the associations interfered with or altered that learning is some way and, certainly, a non-verbal conditioned response would have been preferable. Nevertheless, the OCD group showed a similarly general elevation in performance to controls in the general-PIT test. We also could not find any change in the size of the deficits in the choice tests in those with OCD when we restricted our analyses to participants that overlapped with controls on our conditioning measures (see supplementary results).

Importantly, these deficits were associated with changes in the neural circuits we identified in healthy controls. The most notable effects were patterns of hypoactivity in the lateral OFC (BA47), during the PIT test, and anterior PFC (BA10) extending to the dorsal ACC (BA32) during the outcome devaluation test. These relative changes were accompanied by hypoactivity in striatal targets in caudate and putamen implicating orbitostriatal and prefrontal striatal disconnection in these effects. Interestingly, specific-PIT was also accompanied by hyperactivity relative to controls in the medial OFC (BA11). Although in a slightly more anterior region of BA11 to general PIT, this hyperactivity is consistent with the absence of specific reward predictions, suggesting the OCD participants were responding to the general, reward predictions of the stimuli. If so, this activity was clearly suppressed in the healthy participants, perhaps reflecting the well documented role of lateral OFC (BA47) in the inhibitory regulation of sensation, emotion and cognition for action(41,42). (41,42). Similarly, the deficit in the outcome devaluation test in participants with OCD may also reflect a loss of inhibitory control as a result of the observed hypoactivity in dorsal ACC (BA32), an area often implicated in conflict resolution(43–45) and potentially necessary to resolve the conflict when retrieving the previously and more recently experienced outcome values before and after devaluation(46).

These findings resonate with evidence showing impaired cognitive flexibility in OCD: in adults, in their ability to report the contingency between action and outcome(47,48), and, in adolescents, in tests assessing learning and memory and the ability to use specific stimulus-outcome associations to guide go and no-go discrimination(49). However, the current tests go further in pointing to broader deficits in the emotional and motivational processes that control instrumental performance. Deficits in goal-directed control have been interpreted as suggesting the intrusive thoughts and behavioral compulsions in OCD reflect a shift to an automated or habitual process(23,50). However, the pattern of deficits observed here appear to be more closely associated with dysregulated action control due to specific failures of inhibition during conflict, resulting in the intrusion of actions irrelevant to achieving currently adaptive goals. Previous research has linked anterior medial OFC in particular with the retrieval of specific action-outcome associations and a deficit in this retrieval would necessarily reduce the influence of specific outcome predictions on choice(51). The medial OFC hyperactivity during specific-PIT is consistent with a failure to inhibit alternative actions. Indeed, some time ago Modell and colleagues developed a circuitry model of compulsions in OCD with striking similarities to the circuit thought to support Pavlovian-instrumental transfer(52). They argued that OCD pathology induces dysregulation of a limbic-striatal-thalamic circuit modulating medial OFC resulting in compulsive symptoms, a view that resonates with the medial OFC activity observed during specific PIT in the current study.

Our focus on adolescents to advance an understanding of the causal role of OCD pathophysiology is important given OCD begins during childhood or adolescence in 80% of people(53) and in that group has greater genetic contribution (45–65%) than adult-onset disorder (27–47%)(54). Moreover, premorbid ritualised behavior in early childhood occurs in probands and strong reactions to everyday sensory events are associated with high childhood ritualism(55,56). The current data suggest that these repetitive behaviors may be an early manifestation of an impairment in the motivational control of goal-directed action, presenting a behavioral marker that, combined with a family history of illness, might predict disease onset and indicate early intervention. This is particularly important given that meta-analysis of results from clinically developed behavioral tests often do not show cognitive impairment in children with OCD(57), something that questions whether these impairments are a consequence of the illness or their social and experiential sequelae(58,59), perhaps associated with the fundamental motivational and emotional deficits described here.

## Data Availability

All data produced in the present study are available upon reasonable request to the authors.

## Acknowledgements

The authors thank Laura Bradfield for helpful discussions regarding the content of this paper. This research was supported by a post graduate scholarship from the: National Health and Medical Research Council (NHMRC) of Australia (#1134268) and research funding from the New South Wales Institute of Psychiatry to IEP; and funding from the Australian Research Council #FL990992409 and a NHMRC Senior Investigator Award GNT1175420 to BWB. A preprint of this manuscript has been posted on medRxiv.

## Author contributions

Conception: IEP and BWB. Design: IEP, RWM, SQ, KG, FW, MO, PLH, and BWB. Data acquisition: IEP, RWM, SQ, FW, and MO. Data analysis: IEP, RWM, SQ, and KG. Data interpretation: IEP, RWM, SQ, KG, PLH, and BWB. Drafted or substantively revised the manuscript: IEP, RWM, KG, PLH, and BWB. All authors approved the submitted manuscript and agreed both to be personally accountable for the author’s own contributions and to questions related to the accuracy or integrity of any part of the work.

## Supplementary material

Perkes et al The motivational determinants of human action, their neural bases and the functional impact of OCD

### [1] Full experimental methods including participant instructions, data handling, data analysis and imaging procedures

#### Design

We conducted a case-control cross-sectional study using associative learning paradigms, gold-standard clinical phenotyping, and multimodal magnetic resonance imaging. This study had approval (number 2012/2284) from The University of Sydney Human Research Ethics Committee.

#### Participants

21 healthy adolescents (control group) and 20 adolescents with a lifetime DSM-5 diagnosis of OCD (OCD group) were included in analysis. The sample size was based on power analysis drawn from a similar study in our lab(13) that stipulated a minimum sample size of n=16 to achieve 80% statistical power at an alpha of 0.05. There were no group demographic differences (**Table 1**). Consent or assent was provided by the participant, parent, or both. General inclusion criteria were: (1) age 12 – 18 years at time of testing, (2) no current DSM-5 eating disorder, (3) no DSM-5 intellectual disability, (4) no severe acquired brain injury, (5) no history of central nervous system infection, (6) no current substance use more frequent than once per month, (7) no food allergies, (8) no MRI contraindications (e.g., full dental braces, other metallic implants). Co-morbid psychiatric diagnosis was allowed in the OCD group to improve external validity. Specific inclusion criteria for controls were: (1) no previously diagnosed DSM 5 disorder (past adjustment disorders and past or present elimination disorders were allowed), (2) no lifetime treatment with psychotropic medication, (3) no first-degree relative with OCD. General exclusion criteria were: (1) structural central nervous system abnormalities, (2) > 2 mm head movement during the scan, (3) failure to comprehend or recall the task instructions. Adolescents with OCD were recruited from 107 consecutive presentations (11/03/2008-09/03/2015) to an OCD clinic freely accessible to the public for children and adolescents residing in a geographical area within Sydney, Australia. Having excluded patients outside the age inclusion criterion at time of the study (n=45) or with a diagnosis of intellectual disability (n=2), 60 candidate participants remained. Telephone contact was attempted with 46, 20 of whom declined to participate, 2 had limited English language proficiency, and 3 had MRI contraindications. The remaining 21 attended for testing, one participant was exluded because the semi-structured clinical assessment excluded OCD. A child and adolescent psychiatrist clinically determined caseness. Recruitment of controls occurred through advertisement, convenience, ‘snowball’, and a research volunteer registry.

Telephone screening for inclusion criteria and recruitment was undertaken – refer **Supplementary Table S1**. Participants completed self-report questionnaires that recorded demographics (age, gender, ethnicity, language, and education), medications (agent, dose, and duration), and the Depression Anxiety and Stress Scale [DASS](25). Pre-morbid intelligence was assessed with the Weschler Ranging Assessment Test [WRAT](26).

All participants in both groups were assessed using the Schedule for Affective Disorders and Schizophrenia for School-Aged (K-SADS –PL 2013). OCD symptom measures(27) were completed for the clinical group. To optimize standardization, all assessments were completed a child and adolescent psychiatry registrar (IEP) who interviewed the participant and one or both parents. A child and adolescent psychiatrist (PLH) provided training and supervision of diagnostic interviews. All clinical assessment data was collected within 24 hours of behavioural experiments.

#### Behavioral stimuli, outcomes & equipment

Outcomes consisted of five different sweet or salty foods, commercially named: Arnott’s Chocolate Tiny Teddy® Biscuits 250g, Doritos® Cheese Supreme Corn Chips 114g, Cheezels® Cheese Snacks 114g, Arnott’s BBQ Shapes® 250g, Milk Chocolate M&M’S® 49g.Task design, stimulus presentation and response recording was controlled by PsychoPy© software (v1.82.00) running on a MacBook© (Apple, CA) computer. Visual stimuli during scanning were displayed a projector placed behind the MRI scanner. A Lumina© MRI-compatible two-button response pad (Cedrus©, California) detected responses. Participants viewed a reflection of the projected image (800 × 600 pixels) in a mirror attached to the scanner headcoil.

#### Procedure & setting

Participants abstained from eating for three hours prior to the experiment. Before training, participants were asked “On a scale of 1 to 10, how hungry are you right now?”. Sealed commercial packages of the five foods were opened onto individual plates in front of participants in order to assuage any concerns about contamination (Arnott’s Chocolate Tiny Teddy® Biscuits 250g, Doritos® Cheese Supreme Corn Chips 114 g, Cheezels® Cheese Snacks 114 g, Arnott’s BBQ Shapes® 250 g, Milk Chocolate M&M’S® 49 g). Participants tasted and rated each food on a 7-point Likert scale (“Very Unpleasant” to “Very Pleasant”). Instrumental and Pavlovian conditioning were conducted in an interview room. Participants were verbally asked 6 open questions assessing their knowledge of the instrumental and Pavlovian associations; if an answer was incorrect then the participant was asked “Could it have been something else?”, if all questions were answered correctly then a positive affirmation was given. The Pavlovian-instrumental transfer and outcome devaluation behavioural tests were completed during fMRI data acquisition. Written instructions were shown to participants on the computer monitor. Throughout the task, a virtual ‘snack vending machine’ image was intermittently presented on the screen. Participants learned how to acquire food rewards from this vending machine. Verbal instruction in response to questions from participants was limited to generic responses such as “Tip the machine to learn how to earn the snacks”. To improve reliability, one researcher (IEP) conducted all clinical assessments and behavioural experiments.

#### Behavioral Methods

##### Instrumental conditioning

Left (A1) and right (A2) button presses were reinforced on a variable-ratio schedule (VR5) with a specific food, counterbalanced, dependent upon each participant’s three highest rated foods (**Figure 1**). The plate of snacks (O1) associated with A1 was placed on the desk on the left-hand side of the participant and the plate of snacks (O2) associated with A2 was placed on the right-hand side of the participant. As each outcome was earned, an image of that food appeared on the screen for 1 second and participants were invited to eat one piece of the relevant food. The following instructions were presented on screen: “You can get free snacks from our vending machine. Tip the machine with the left or right arrows. Learn how to get the different snacks. Press any key to begin”. After every third outcome participants were asked: “Which direction did you tilt to get (the outcome)”. Feedback was provided (“correct” or “Oops! That was wrong”). Instrumental conditioning ceased after a participant registered six consecutive correct answers.

##### Pavlovian conditioning

Prior to the start of conditioning the button box was removed. The three plates holding all three food rewards (O1, O2, O3) involved in Pavlovian conditioning were placed on the desk. Four stimuli (S1, S2, S3, S4) were paired with four outcomes (O1, O2, O3, Ø) (**Figure 1**). Two stimuli (S1, S2) were paired with two outcomes from instrumental conditioning; i.e., ensuring that S1-O1 and S2-O2 were distinct pairs. One of the stimuli (S3) was paired with an outcome (O3) that was not included in instrumental training stage. The fourth and final stimuli (S4) was paired with the word ‘EMPTY’ indicating that no food was available. The following instructions were presented on screen: “The vending machine cannot be tipped now. But, free snacks will sometimes fall out. Coloured lights will appear on the machine before a snack falls out. Watch the lights and learn which snack will fall out. Questions will test what you learn”. Stimuli were presented for 5 seconds, after which the image of the food outcome appeared beneath the stimuli (coloured vending machine) for 1 second — a total of six seconds. The inter-trial-interval (ITI) was 10 (+/-5) s and during the ITI the vending machine was shown without either stimuli (colour) or outcome (food). After every block of four stimuli-outcome trials a multiple-choice question “Which snack will fall out?” appeared on the screen with a stimulus (coloured vending machine), if participants answered this question correctly then they were invited to eat one piece of the relevant outcomes. Pavlovian conditioning ceased after a participant registered six consecutive correct answers.

##### Pavlovian-instrumental transfer test

For this test the button box was returned and the four stimuli were presented individually for 6 seconds every 18 seconds (0-4 second random jitter). Each stimulus was presented 12 times in random order. Participants were able to tilt the vending machine during stimulus presentation and when the vending machine was unlit during the intertrial interval, providing an active baseline measure (**Figure 1**). This transfer phase was conducted in extinction, i.e., no outcomes, to ensure that responding was not influenced by change in the incidence of outcome delivery during the test. The following instructions were presented on screen: “The vending machine will now sometimes give free snacks. You will see coloured lights on the machine again. You can tip the machine at any time. No snacks will appear on the screen, but the snacks you earn will be recorded. Remember what you learned before to get all the snacks that you want!” Pavlovian-instrumental transfer data for one participant from each group was missing due to a data recording error, leaving OCD *n*=19 and controls *n*=20.

##### Outcome devaluation procedure and test (**Figure 2**)

Participants were first shown the following statement on screen: “Now you’ll see what has happened to one of the snacks!” After this statement they were shown a 4-minute video of cockroaches crawling on one of the foods (counterbalanced between O1 and O2) they had learned to earn during instrumental conditioning. After the video presentation the following instructions were presented on screen: “You return to the vending machine you saw before. You can tip the machine at any time. No coloured lights or snacks will appear, but a tally will be kept of the snacks you get. Get all the snacks that you want!” The blank vending machine then appeared for 30 trials of 12 seconds each. Before each trial, a fixation cross was presented for 18 (±6) seconds. Participants could tilt the machine or fixation cross at any time. No outcomes were presented during the devaluation test.

##### Recall test (post-test questionnaires)

After the devaluation test, whilst still in the scanner, participants rated the desirability of O1 and O2 on a scale of Likert scale 1 to 7. After exiting the scanner participants re-completed the self-report hunger and food pleasantness scales that were first completed at the start of the behavioural experiments. They also completed a self-report six-item multiple-choice test of their ability to recall of the instrumental (e.g., ‘What snack was associated with the LEFT key?’) and Pavlovian (e.g., ‘What snack was associated with the BLUE light?’) contingencies.

#### Imaging methods

Scanning occurred in a 3T GE Discovery with a 32-channel head coil (GE Healthcare, UK).

A T1-weighted high-resolution was acquired for each participant for registration and anatomical screening: 7200-msec repetition time; 2700-msec echo time; 176 slices in the sagittal plane; 1-mm slice thickness (no gap); 256-mm field of view; and 256 × 256 matrix.

We acquired 300 T2*-weighted whole-brain echo planar images with a 2910-msec repetition time (TR); 20-msec echo time; 90-degree flip angle; 240-mm field of view; and 128 × 128 matrix with SENSE (Sensitivity Encoding). Each volume consisted of 52 axial slices (2-mm thick) with a 0.2-mm gap. Whole brain diffusion-weighted images were acquired using an echo planar imaging sequence with the following parameters: TR=8250ms; TE=85ms; number of slices=55 thickness=2mm-thick axial slices; matrix size, 128 × 128; in-plane resolution, 1.8 × 1.8mm^2^; 69 gradient directions. Eight images without gradient loading (B0 s.mm-2) were acquired prior to the acquisition of 69 images with uniform gradient loading (B0=1000s.mm-2).

#### Data Analysis

Descriptive statistics were calculated and two-tailed t-tests were used for continuous variables and chai-squared for categorical variables. Selective serotonin reuptake inhibitor (SSRI) medication doses were standardized mg equivalents for fMRI analysis.

##### Pavlovian-instrumental transfer (predicted value)

Outcome-specific PIT was determined by a comparison of the rate of the ‘same’ action and the ‘different’ action during the S1 and S2 stimulus trials. During S1 trials, the ‘same’ action was A1 and the ‘different’ action was A2.

During S2 trials, the reverse was true: the ‘same’ action was A2 and the ‘different’ action was A1. The number of same and different actions was calculated per trial for each participant. For group differences in behaviour, the average ‘same’ and ‘different’ action rates were summarised per person and included in a 2 (group) x 2 (action) mixed ANOVA, where the interaction determined whether specific PIT was aberrant in OCD. For the fMRI analysis, the difference between the rate of the ‘same’ action less the ‘different’ action per trial was calculated, for each person. This was used as a parametric task regressor for specific PIT in the fMRI analysis (described below). For the tractography correlation analysis, this average difference was included as a covariate to determine tract weights related to specific PIT.

**General transfer** was determined by comparing the rate of actions during S3 and S4 stimuli. The action rates (aggregate button-presses) were calculated per trial for each person. For group differences in behaviour, the average S3 and S4 response rates were summarised per person and included in a 2 (group) by 2 (stimulus) mixed ANOVA, where the interaction determined whether general-transfer behaviour was aberrant in OCD. For the fMRI analysis, the vector of these rates was used as a parametric task regressor for general PIT.

Baseline rates were calculated as the total number of button presses per second during presentation of the ‘blank’ vending machine, and average group differences were tested with a 2-sample t-test.

##### Outcome devaluation (experienced value)

The effect of the outcome devaluation procedure was determined by the change in food preference ratings across the battery of experiments, i.e., Δ value = pre-rating – post-rating. The interaction in a 2 (group) x 2 (pre-post) mixed ANOVA on the change scores indicated whether desire was aberrant in OCD. The effect of devaluation on behaviour was determined by the rate of actions for the still-valued food was calculated per trial, for each person. Group differences in goal-directed behaviour were tested in a 2 (group) x 2 (action) x 5 (trial bin) mixed ANOVA on the average still-valued and devalued action rates per person, where a significant interaction between group and action (valued vs devalued) indicated aberrant goal-directed behaviour in OCD. Valued-devalued action rates per trial were included as a parametric task regressor in the fMRI analysis. The average difference in valued action rates less devalued rates for each person was also included in the tractography analyses to determine tract weights related to goal-directed behaviour.

##### Magnetic Resonance Imaging – Functional (fMRI)

The data from each test were analyzed separately using SPM8 (Wellcome Department of Imaging Neuroscience). Structural images were manually inspected for anatomical abnormalities and co-registered to the mean functional image. Functional images were realigned, slice-time corrected, normalized to the Montreal Neurological Institute (MNI) template space, interpolated to 2 × 2 × 2 mm voxels and smoothed with a Gaussian filter (8-mm full width-half maximum). To correct for movement on image analysis we distinguished inter-subject motion and task-correlated motion. Subject motion can produce image artifacts (e.g., banding) which increases the error term in the statistical model and reduces the likelihood of correctly detecting a significant effect. To address this, we screened each run after movement correction and normalization (i.e., post-processing) for image artefacts using the Artifact Detection Tool from Susan Whitfield-Gabrielli (web.mit.edu/swg/software.htm). For each participant, outlier images were identified using the scan-to-scan differences in movement (mm) and rotation (degree) with default thresholds of 2 mm and 0.2 degrees, respectively. These points were used to construct an outlier regressor for each individual to be added as a covariate in the first-level analysis (see below). This resulted in the exclusion of 2.6 percent of data in the OCD group (highest percent from any single participant was 21.3 percent) while 0.4 percent of data was excluded among the control group. Using the same Artifact Detection Tool, we also manually screened each run for task-correlated motion which will increase the false positive error rate. There were no substantial correlations with any task regressor in our sample, mean r=0.08 (highest r=.19). The six movement regressors from realignment were also included as regressors-of-no-interest in each GLM (described below).

The fMRI analyses were conducted in a two-level manner, where the first-level specified a general linear model (GLM) for each participant, and the second-level included the first-level parameters as random effects. The first-level GLM for the Pavlovian-instrumental transfer test modelled conditioned stimuli as a boxcar function with separate regressors for specific-(S1, S2) and general-(S3, S4) stimuli. We modelled response times as stick functions in a separate regressor of no interest. Following Prevost et al 2012, a parametric regressor modulated the S1 and S2 stimulus blocks by a vector of the difference between ‘same’ and ‘different’ action rates per stimulus as the trial-wise task regressor for specific transfer. The S3 and S4 stimulus blocks were parametrically modulated by the vector of total response rates per stimulus, which served as the (trial-wise) task regressor for general transfer. The first-level GLM for the devaluation test included trials as a boxcar function and a regressor-of-no-interest modelling response times as a stick function. A parametric regressor modulated the trial blocks by the number of valued responses over devalued responses, which served as the (trial-wise) task regressor for choices driven by experienced value (following Morris et al 2015). Each task regressor was convolved with the canonical haemodynamic response function (after high-pass filtering with a cut-off of 128 s to remove drifts within sessions).

The resulting parameter estimates (betas) for the task regressors were entered into second-level *t*-tests in SPM8 to generate population-level effect statistics. BOLD activity tracking each task regressor were tested in planned whole-brain one-sample SPM *t*-tests of betas from healthy adolescents, while aberrant BOLD activity in OCD was tested in planned whole-brain two-sample SPM *t*-tests of betas from both groups. Significant regions in each whole-brain analysis, exceeding a voxel level false-discovery rate FDR *q*=.05 are reported here (clustersize threshold *k=*5). Follow-up region-of-interest (ROI) analyses comparing groups in regions implicated by the task regressor in health adolescents were also performed when the planned group comparison was null, and results exceeding a small-volume corrected family-wise error rate *p*=.05 are reported. We also performed ROI analyses for correlations with obsessions (e.g., contamination, disgust, or symmetry) or compulsions, training performance, age, WRAT scores (IQ proxy), handedness, and SSRI medication dose. Significant regions were manually verified using the Atlas of the Human Brain.

##### Diffusion Imaging and Tractography

Diffusion data was first eddy-current corrected using FMRIB Diffusion Toolbox to align all images to a reference b0 image and linearly transform them, brains were extracted, and diffusion tensors fitted. Diffusion probabilistic tractography was then performed using the FDT Diffusion Toolbox. We determined seed masks using clusters of significant activation from the preceding fMRI analysis. For each participant, tractography was performed from every voxel within the seed mask to build up a connectivity distribution. We fitted a three-fibre orientation diffusion model to estimate probability distributions on the direction of fibre populations at each brain voxel in the diffusion space of each participant. To interpret the probabilistic tractography in standard space, we used standard-to-diffusion matrices and the corresponding inversed matrices. We generated 5000 samples from each seed voxel with a curvature threshold of 0.2 and no waypoint or termination masks. Tracking occurred in diffusion space, with results transformed back to MNI space. To visualize tracts efferent and afferent to the seed mask, individual participant 3D files were thresholded to the top 0.02% of tracts and binarized, before being concatenated into a 4D file. This showed the average connectivity, across all participants, for each seed region. FSL (FMRIB Software Library) tools (www.fmrib.ox.ac.uk) were used in all diffusion analyses (version 5.0.1).

We tested for group differences in the estimated strength of tracts efferent and afferent to our seed regions using nonparametric voxelwise statistical testing, and assessed the relationship between tract strength and the behavioural covariates (average rate of specific transfer from the task regressor, and the difference between valued and devalued press rates in the first minute of test, for PIT and outcome devaluation, respectively) with the tract values at each voxel, independently for each of the seed regions.

After group comparisons and voxelwise correlations against the behavioural regressors, the model fit was tested by permutation testing (FSL Randomize), using 25 000 random permutations. Threshold-free cluster enhancement (TFCE) was used to boost signal in areas that exhibit spatial clustering. To protect against false positives, we restricted the analysis to those voxels in which at least half of the participants (n=19) had tracts from the seed mask. In addition, only clusters of at least 20 contiguous voxels are reported.

Resulting statistical maps were thresholded at p=0.05 family-wise error corrected (FWE). A significant relationship between white matter tractography values and behavioural regressors at a particular voxel implies variable white matter architecture between (some part of) the seed region and the voxel in question.

### Supplementary Results

#### The influence of Pavlovian conditioning on specific PIT and the deficit in OCD

To investigate whether the degree of Pavlovian conditioning influenced specific PIT, we conducted a post-hoc analysis of participants who remembered all Pavlovian and instrumental contingencies at the end of the experiment excluding participants with a post-test memory score less than 100 percent (2 controls and 5 OCD) leaving OCD *n*=14 and controls *n*=18 in each group for this analysis. The mean response rates among these subgroups of ‘complete learners’ confirmed that the marked deficit in specific transfer remained in the OCD relative to the control group (**see Figure 2A inset**, group interaction *F*_1,30_=5.23, *p*=.029, 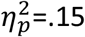), alongside intact stimulus-elicited arousal during general transfer (**Figure 2B inset**, main effect of cue *F*_1,30_ =9.93, *p*=.004, 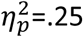; group interaction *F* < 1).

#### Covariate analyses of specific PIT test

An analysis including age or WRAT score as a covariate did not change the pattern of results between groups (e.g., largest group difference in medial OFC remained: BA11: MNI: -4,50,-20, *F*_2,36_=32.30, pFDR < .001 after controlling for age). Likewise, excluding three left-handed OCD participants did not alter the pattern of significant results between groups (e.g., largest group difference remained in left MOrG (BA11: MNI: -6,46,-24, *F*_2,34_=32.47, pFDR < .001). An analysis limited to the OCD group and including antidepressant dose (fluoxetine equivalent dose) as a covariate did not reveal any significant effects of medication (pFDR=.873).

#### Covariate analyses of outcome devaluation

The majority of the OCD group had obsessions with contamination or disgust obsessions (*n*=16; 80%). The effect of revaluation in this subgroup was significant (*p*=.007, Cohen’s *d*=0.77, corrected for dependence between means) and the effect size was similar or larger than that among the total OCD group (i.e., Cohen’s *d*=0.52, corrected). However, the correlation between symptom severity and choices or ratings after revaluation in this subgroup were small and non-significant (*r*s < .361).

#### Covariate analyses of the choice devaluation effect

Analysis including age or WRAT score as a covariate did not change the significant group difference in the correlation with Anterior PFC activity: BA10: MNI: 2,50,10, *F*_2,38_ = 9.04, pFWE = .035, svc, after controlling for age whereas *F*_2,38_=9.82, pFWE=.024, svc, after controlling for WRAT. Likewise, excluding three left-handed OCD participants did not alter the significant result (MNI: 2,50,10, *F*_2,36_=8.75, pFWE=.044, svc). A further analysis limited to the OCD group and including antidepressant dose (fluoxetine equivalent dose) as a covariate again failed to reveal any significant effects of medication (pFDR=.279).

#### Tractography

##### Specific PIT

Although the tractography analysis supported the cortical network implicated in the influence of predicted value, we found no significant differences in tract strength when we used the largest between-group differences in the fMRI results for specific transfer as seed regions. **Figure S1A** shows a significant negative correlation with the average rate of specific transfer and the tract strength between the left lateral OFC and the middle frontal gyrus (peak voxel -28 10 24, pFWE=.047, cluster size 286 voxels). In other words, across both groups, the stronger the OFC–MFG tract connection the weaker the influence of predicted values on choice during outcome-specific PIT. **Figures S1B and S1C** show the raw tractography thresholded at the top 0.02% of efferent and afferent tracts from the medial OFC and right lateral OFC, respectively, across all participants.

#### Outcome Devaluation

Using the hypoactive anterior PFC/dorsal ACC region identified in our analysis of OFC vs. HA in outcome devaluation as a seed region, tractography analysis compared afferent and efferent tracts between groups to investigate neural disconnection as a contributing factor to task performance. This analysis revealed a lower tract strength in the projection to the caudate nucleus (peak MNI coordinates: 6,18,-6; cluster size: 56 voxels) in adolescents with OCD relative to healthy controls (pFWE=.008; **Supplementary Figure S2A**). See **Supplementary Figure S2A** and **Supplementary Figure S2B**.

**Table S1.**
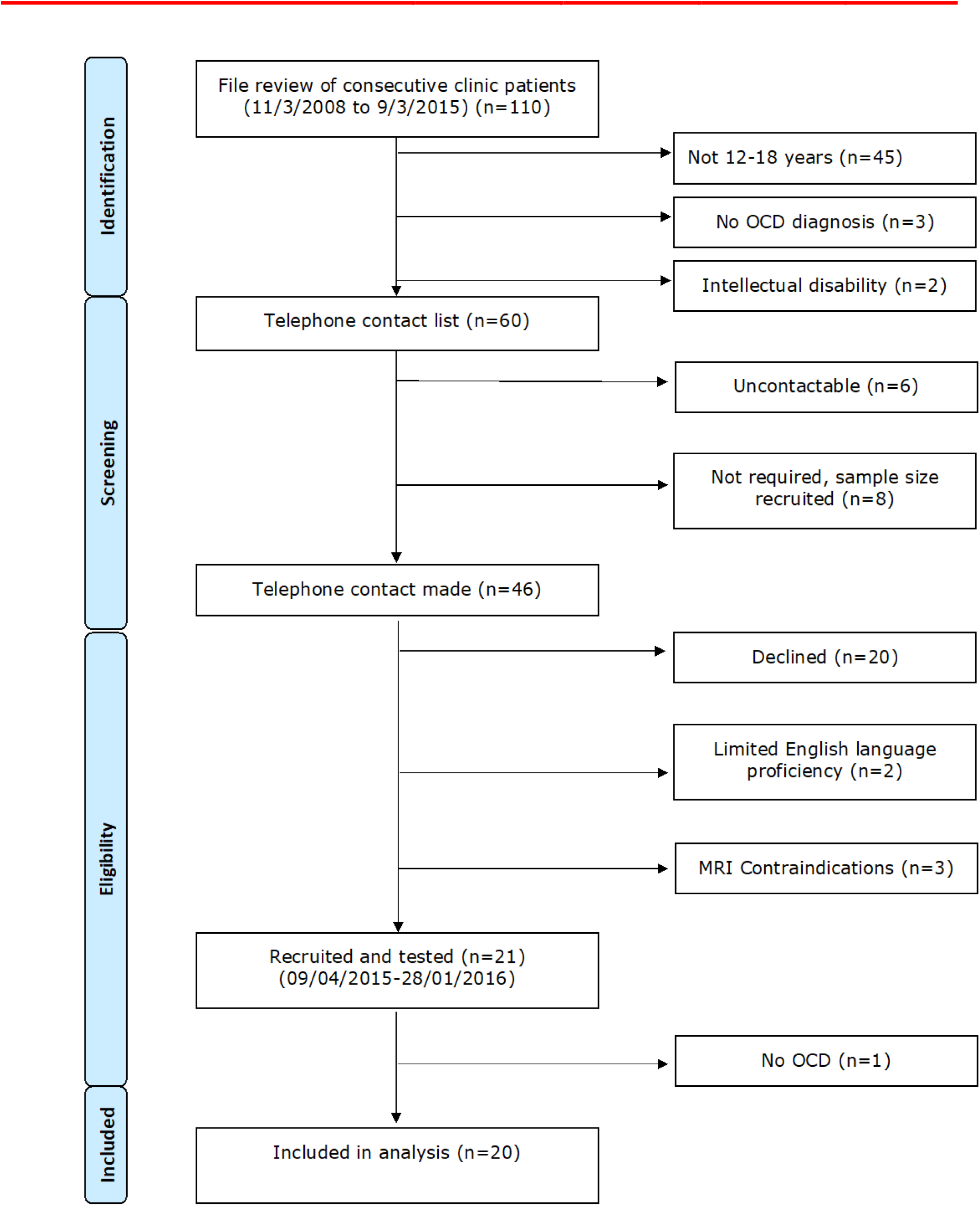
Recruitment flowchart

**Figure S1:**
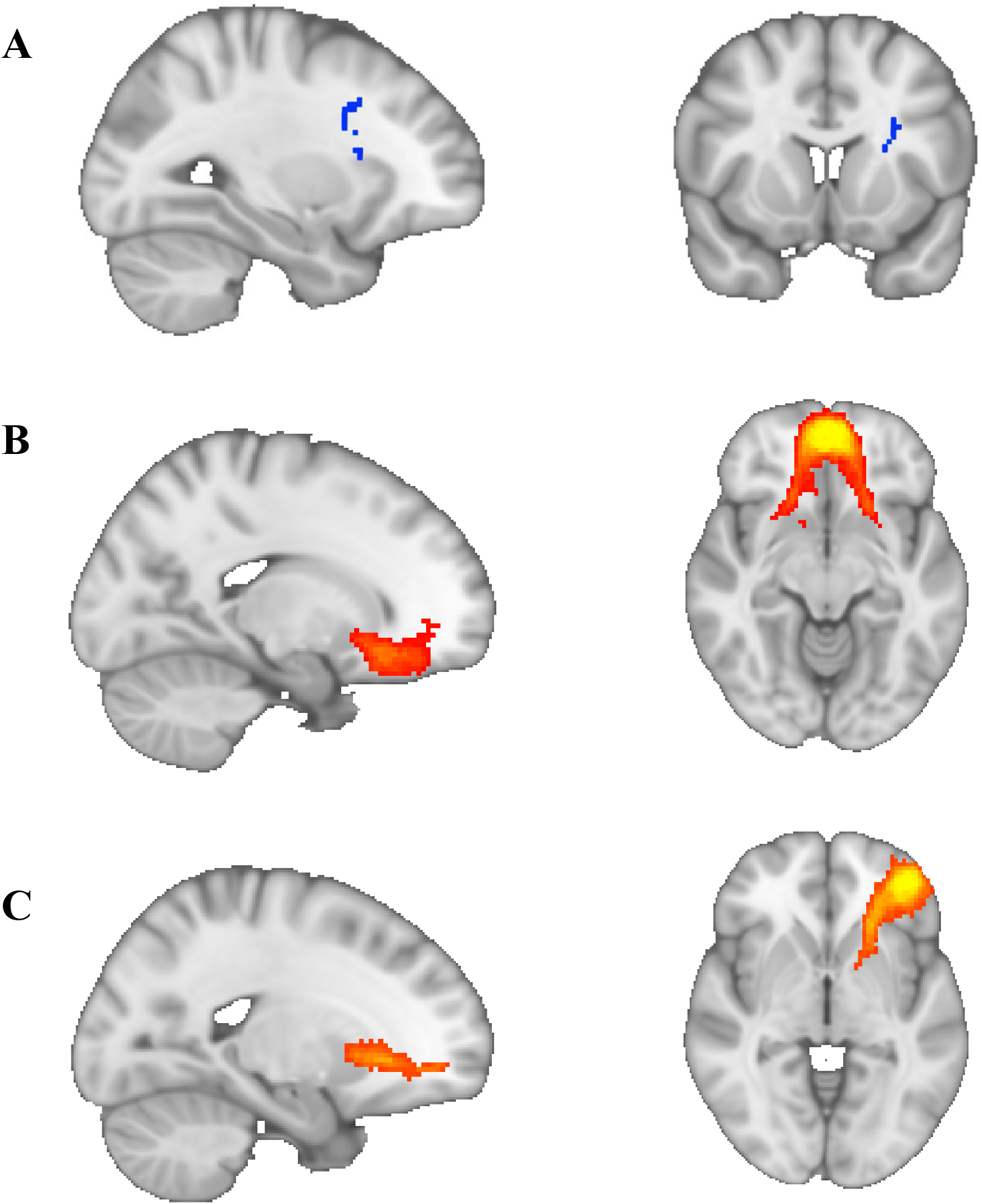
Tractography in Specific PIT test. (**A**) We found a significant negative correlation between tract strength between the left lateral OFC and the middle frontal gyrus and the average rate of specific transfer (peak voxel -28 10 24, cluster size: 286 voxels). Raw tractography thresholded at the top 0.02% of efferent and afferent tracts from (**B**) medial OFC, and (**C**) right lateral OFC across all participants. There were no between-group differences in tract strength with any of the seed regions tested.

**Figure S2:**
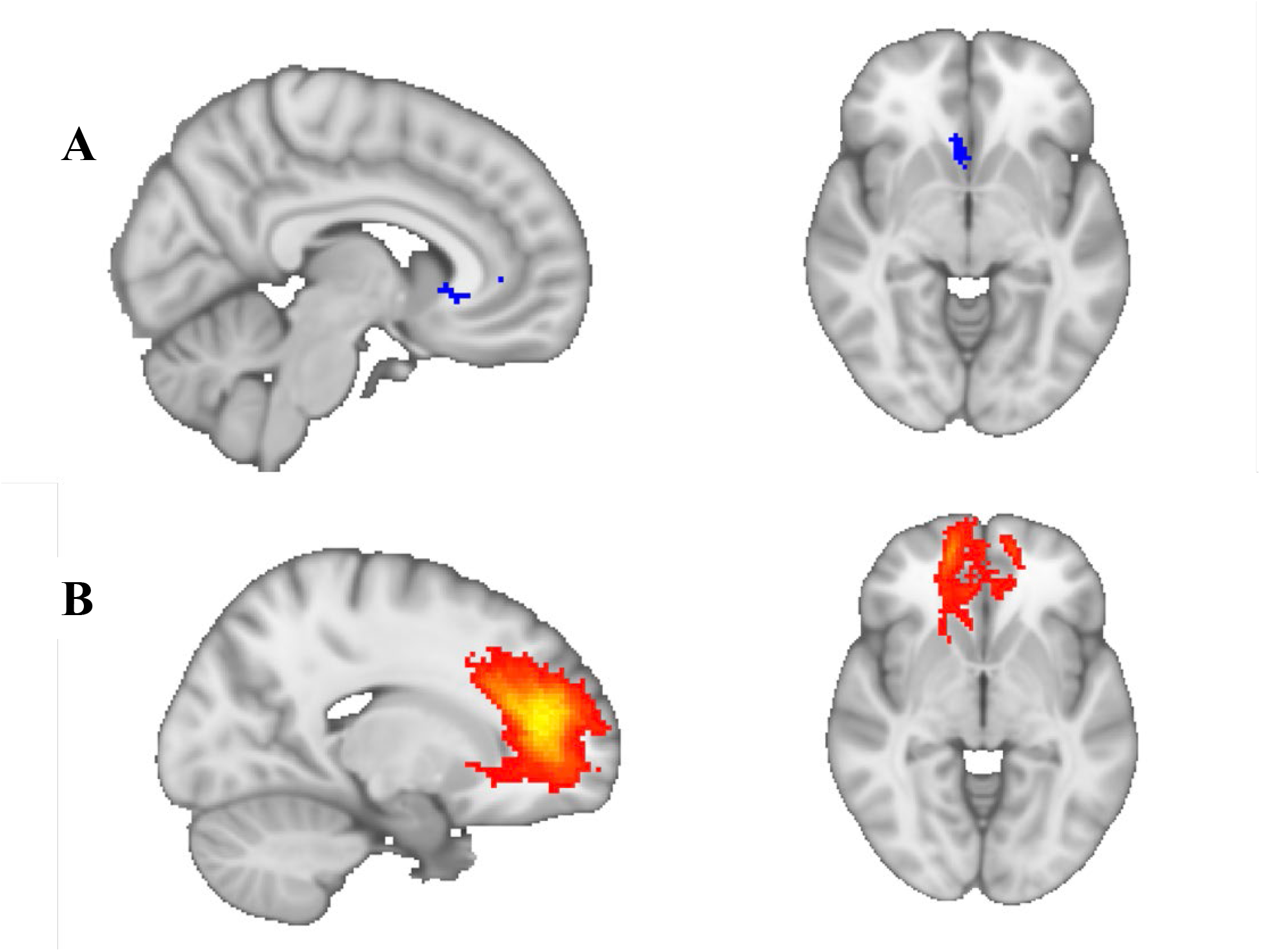
tractography for Outcome Devaluation Test. (**A**) We found significantly lower anterior PFC/anterior cingulate complex-to-head of caudate (peak MNI coordinates: 6, 18, -6, cluster size: 56 voxels) tract strength in adolescents with OCD relative to healthy controls. (**B**) The voxels with strongest connectivity, across all participants, to the BA10/32 seed mask (top 0.02% of tracts sent from this seed mask).

